# The increasing trend of Non-Communicable Diseases among young adults may invite multiple public health challenges in the future

**DOI:** 10.1101/2024.07.30.24311217

**Authors:** Ashis Biswas, Sharmistha Roy

## Abstract

**Objective:** To compare the burden of communicable and non-communicable diseases (NCDs) in the USA and globally, particularly among 15-45 years old.

**Methods:** The imported data from the WHO Mortality Database portal was analyzed by SPSS.

**Results:** The study shows the percentage of death from NCDs out of total death in the USA in 2010 and 2019 were 86.7% and 86.8% which are higher than global rates of 78.1% and 80.2%. Despite a similar death percentage out of total death, the death rate due to NCDs increased from 601.04/100,000 (**2010**) to 687.99/100,000 (**2019**) and 790.29/100,000 (**2020**) globally; and from 693.60/100,000 (**2010**) to 753.35/100000 (2019) and 795.78/100000 (2020) in the USA. On the other hand, the death rate from communicable diseases were 56.57/100000, 55.52/100000, and 129.47/100000 globally; and 44.63/100000, 43.20/100000, and 151.99/100000 in the USA in the respective years. Young adults of 15-45 years show a higher NCD death rate in the USA with an increasing trend.

**Conclusion:** The USA has a higher NCDs dependent mortality rate relative to the global mortality rate, particularly among the 15-45 years age group. During the COVID-19 pandemic, the relative raising in the communicable disease death rate indicates an interplay between communicable and NCDs.

## Introduction

Death is a natural destination of any living creature including humans. People are trying to ensure a happy and healthy life journey with an increased life span. This aim derived human civilization towards industrialization and this changed people’s lifestyles, food habits, and healthcare. At this point, it seems the transformation is appearing as suicidal. Once the fight was against pathogenic diseases. Now we are mainly fighting against the diseases developing inside our bodies.

Communicable and non-communicable diseases (NCDs) are two major categories of health conditions that affect people worldwide (1). The burden of the disease varies greatly across the globe, with some regions experiencing high rates of communicable diseases while others face rising rates of non-communicable diseases (2,3). The USA is no exception to this trend, and understanding the patterns and trends in disease-related mortality is crucial for improving public health policy and practice (4). This study aimed to compare death rates from communicable and NCDs in the USA to a cumulative global figure using data collected from the World Health Organization (WHO) over the last couple of decades. All races, sex, and age groups were included in the analysis to ensure a comprehensive view of the issue.

Communicable diseases, such as malaria, tuberculosis, and HIV/AIDS, have long been a major global health concern, particularly in low- and middle-income countries (1). According to the World Health Organization (WHO), communicable diseases are responsible for over 9 million deaths annually, with pneumonia, tuberculosis, and diarrheal diseases being among the most common causes of mortality (5).

Noncommunicable diseases (NCDs), such as heart disease, cancer, and diabetes, are now the leading cause of death worldwide, and in many countries, they have surpassed communicable diseases as the primary cause of mortality (3). According to the CDC, non-communicable diseases are responsible for over 7 out of every 10 deaths in the United States, with heart disease and cancer being the leading causes of mortality (6). Globally, NCDs are responsible for over 70% of deaths and are a significant contributor to the burden of disease in low- and middle-income countries (4). According to the WHO, non-communicable diseases are projected to account for over 75% of all deaths globally by 2030 (5). However, the picture is more complex in the USA, where both communicable and NCDs are major public health concerns (4).

Some key factors have been found to contribute to death rates from communicable and non-communicable diseases in the USA and globally over the last ten years. Lower socioeconomic status has been associated with higher death rates from both communicable and non-communicable diseases, likely due to factors such as limited access to healthcare, unhealthy living conditions, and greater exposure to infectious agents(4). Certain lifestyle factors, such as poor diet, physical inactivity, smoking, and excessive alcohol consumption, have been linked to higher death rates from non-communicable diseases, such as cardiovascular disease and cancer (7). Exposure to environmental hazards, such as air pollution, can contribute to higher death rates from both communicable and non-communicable diseases (8,9). In addition, as the global population ages, the burden of non-communicable diseases is expected to increase, leading to higher death rates (10). Limited access to healthcare, including preventative services and treatments, can contribute to higher death rates from both communicable and non-communicable diseases (10–13).

Understanding the patterns and trends in death rates from communicable and NCDs is important for public health officials, policymakers, and researchers (14,15). This study will provide a comparative analysis of mortality rates from these diseases in the USA and other countries, using data from the World Health Organization. By including all races, sex, and age groups in the analysis, this study aims to shed light on differences and similarities in the burden of disease across populations. Ultimately, the findings of this study could inform public health policies and interventions aimed at reducing mortality rates from communicable and non-communicable diseases.

## Methodology

We collected the data from the mortality database of the World Health Organization (WHO) and we used SPSS to analyze the data and to create statistical figures. We use the mortality database for both Non-Communicable Diseases (NCD) and Communicable Diseases for comparative studies between the Global and the USA.

## Result

Non-communicable diseases contribute to most global death and are increasing over time while the emergence of COVID-19 has increased the contribution to the percentage of death specific to communicable out of total death more than two times globally including in the USA. To analyze the global disease burden along with the USA, we have used a mortality database of non-communicable diseases and a mortality database of communicable diseases from the World Health Organization’s mortality database portal.

The global burden of Non-Communicable Diseases (NCD) contributes most to the total burden of global health and it is in an increasing pattern in the last few decades. In figure1 we showed the comparison of the Contribution of Percentage of NCD and communicable diseases out of total deaths between Global and the USA. NCD contributed to more than 76% of global death between the years 2000 to 2019 whereas the contribution of communicable diseases varied between 6-10% except for COVID-19 emergence in 2020 where the percentage was above 13% and NCD contribution dropped to around 76.4%. In the USA the contribution of NCD to the total death was more than 85% between the years 2000 to 2019 with an increasing percentage and the communicable diseases percentage going down from 6.13% to around 5%. In the year 2020, due to COVID-19 the communicable diseases contribution raised to 14.9% and the NCD contribution dropped to 77.8%.

**Figure 1:**
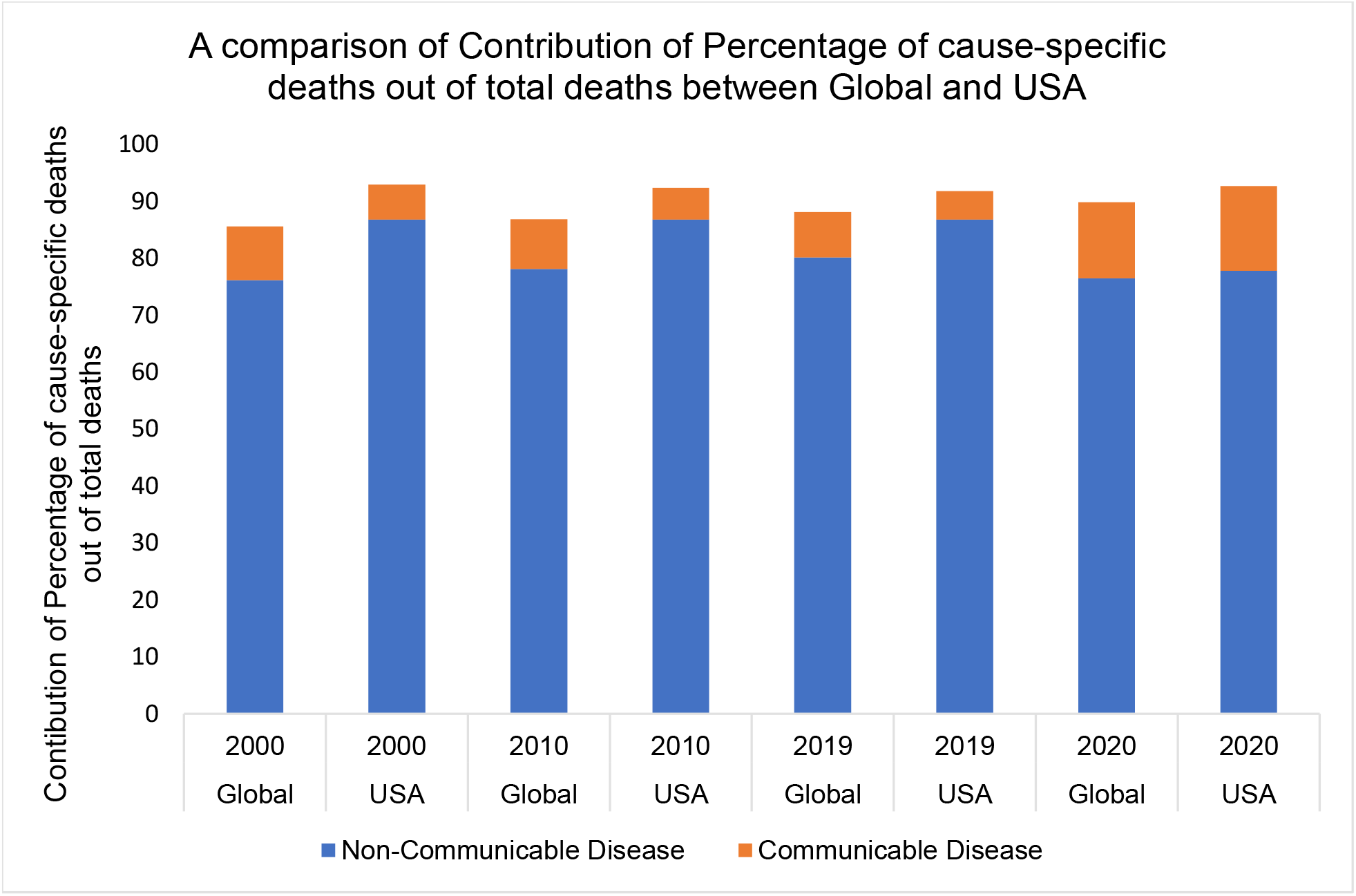
A comparison of the Contribution of Percentage of cause-specific deaths out of total deaths between Global and the USA. Non-communicable diseases contribute to the most death both globally and in the USA and the percentage is increasing. In the year 2020 the percentage of communicable disease increases both globally and in the USA which reflects the emergence of COVID-19.

The USA has a higher death rate from NCD most of the global death is due to NCD. Despite the reduction in percentage contribution to the total death by NCD, the death rate has increased during COVID-19 in 2020 compared to 2019, both globally and locally in the USA. The death rate is higher in the USA during and before the COVID-19 emergence.

In figure 2A, despite the decrease in the percentage of contribution of total death by NCD, in 2010 the death rate from NCD was 601.04 per 100,000 which increased to 687.99 per 100,000 in 2019 and in 2020 during the COVID-19 pandemic, it reached 790.29 per 100,000. On the other hand, in the USA, the death rate in 2010 was 693.60 per 100,000 and went up to 753.35 per 100,00 in 2019, and COVID-19 triggered up to 795.78 per 100,00 in 2020. Figure 2B, shows the death rate due to communicable diseases under the same settings as figure 2A. The death rate from communicable diseases was 56.57 per 100,000, 55.52 per 100,000, and 129.47 per 100000 globally and 44.63 per 100,000, 43.20 per 100,000; and 151.99 per 100,000 in the USA respectively.

**Figure 2:**
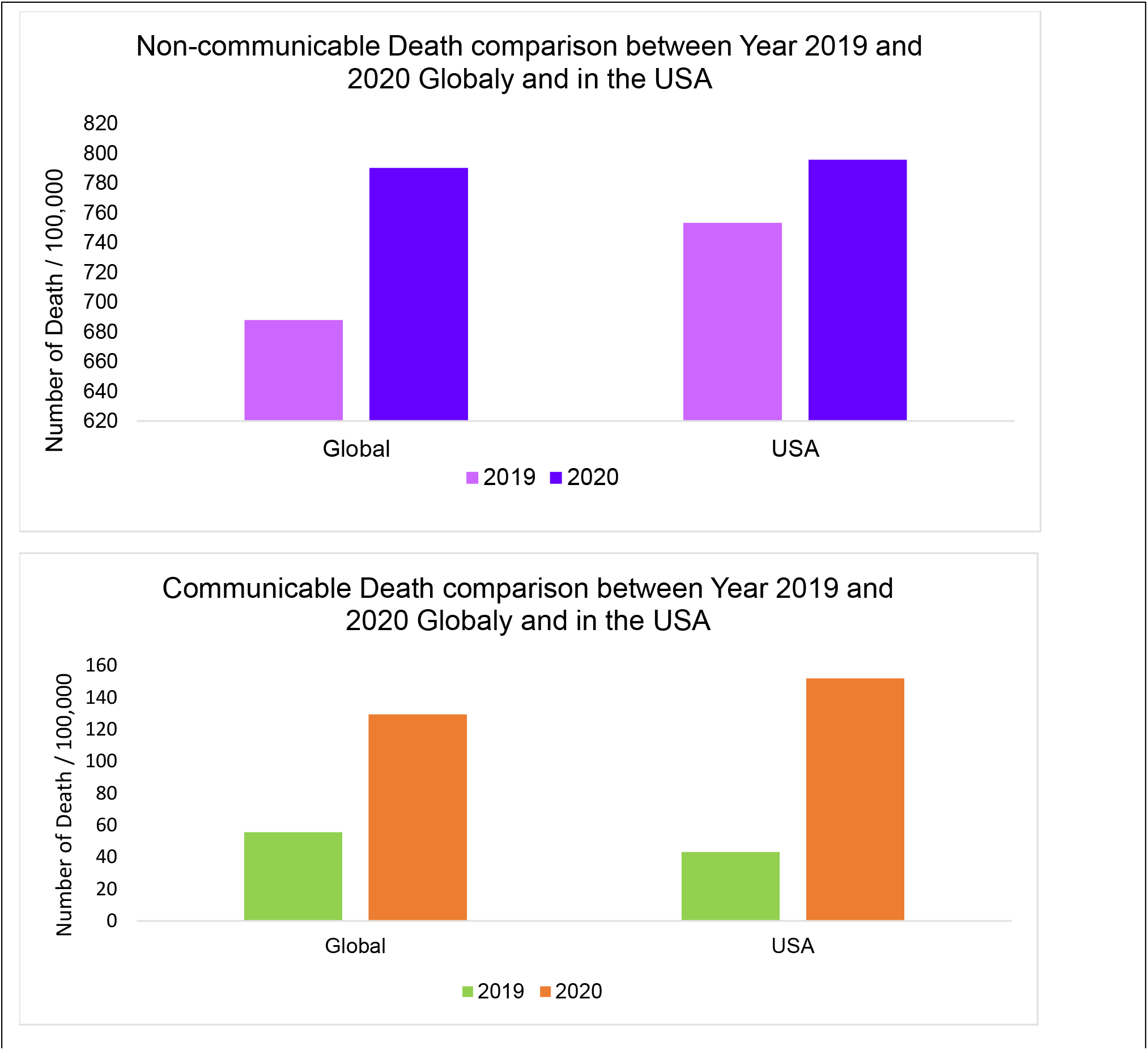
**A**. A comparison of the Death rate due to non-communicable diseases between the Year 2019 and 2020 globally and in the USA. Both globally and in the USA, the non-communicable death rate has increased with the emergence of COVID-19. **B**. Comparison of the Death rate due to communicable diseases between the years 2019 and 2020 globally and in the USA. Both globally and in the USA, the communicable death rate has drastically increased with the emergence of COVID-19.

The mortality rate (per 100,000) due to NCD among the age groups from 15-44 years in Figure 3A shows a comparison between globally and in the USA in the year 2010, 2019, and 2020. From the year 2010, the death rate due to NCD is increasing continuously in the USA among these age groups while globally the rate is consistent.

**Figure 3:**
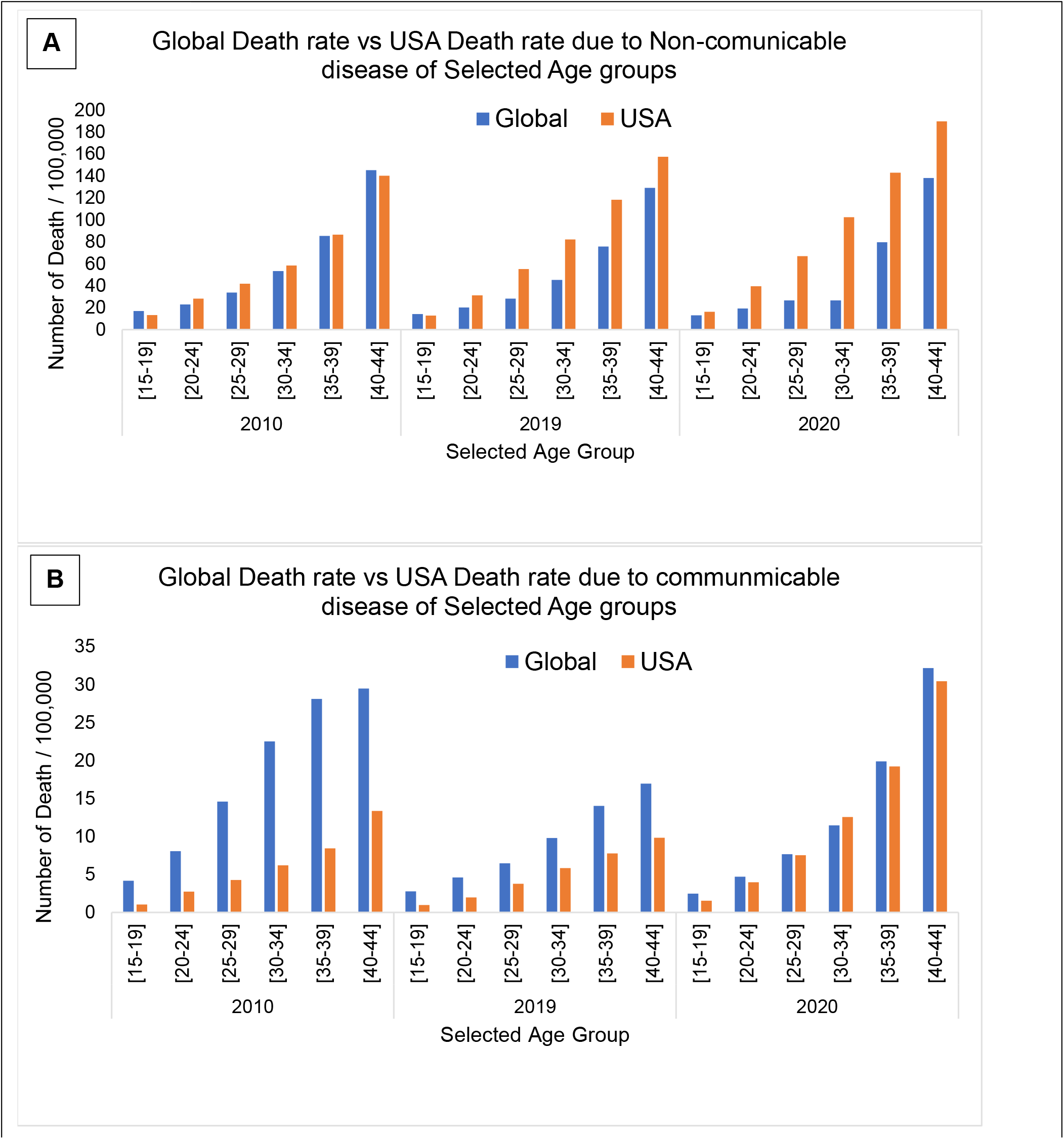
**A**. A comparison between the global death rate and the USA death rate from non-communicable diseases of selected age groups. The death rate increased in most of the listed age groups in 2019 & 2020 compared to 2010 in the USA while globally the death rate is stable. **B**. A comparison between the global death rate and the USA death rate from communicable diseases of selected age groups.

Figure 3B shows a comparison of the death rate (per 100,000) due to communicable diseases among the selected age groups between 15-44 years was higher globally in 2010 and was improving until COVID-19 hits in 2020. In the USA the death rate was lower and went further down in 2019. In 2020, during the COVID-19 pandemic, the death rate globally and in the USA steeply increase and became almost equal among the selected age groups.

## Discussion

To the Global burden of disease (GBD), non-communicable diseases (NCD) contribute the most irrespective of developed, developing, or under-developed countries, and the contribution of Communicable diseases next. In our analysis, we showed the comparison between the global and the USA burden of disease to show how development may impact on contribution to the burden of diseases. The USA features a mini world by providing a home to the most multicultural communities and possessing most ethnic groups. Thus, the statistics of the disease burden of the USA may feature as a global miniature of a developed state.

Analysis of figure 1 indicates that more than 75% of the global disease burden comes from non-communicable diseases (NCD) and in the USA the percentage is higher. During the COVID-19 pandemic in 2020, the percentage of contribution to total death decreased globally, and in the USA does not mean that the total death or death rate due to these conditions was less. In fact, both globally and in the USA the death rate due to NCD went up compared to the previous year (figure 2A). The percentage of death out of total death due to communicable diseases increased during COVID-19 in 2020 globally and in the USA also reflected in the death rate compared in figure 2B. These results, especially for the USA, reflect a synergistic impact of communicable and NCDs on each other leading to a higher rate of death.

Non-communicable diseases require a long period for a phenotype and these diseases are mostly observed among the old age population suggesting the underline reason for the higher death rate from NCD among old ages. But the alarming result is that in the USA, the death rate from NCD is getting higher in the young-age population in a continuous manner in the past decade (figure 3A). These age groups of the population are more important to address as this age is the center of the human life cycle. First, global development depends on these groups of the population. Second, both children and elder citizens in a family depend on this group of family members. Third and the most important is that these age groups of the population are the parents of the progeny. There is a high chance of having NCD among the progeny from the parents with NCDs. From the comparison of NCD-dependent global death rates of this age group was similar to the death rates in the USA in 2010 and science then the rate was increasing in the USA (figure 3A). On the other hand, the rate was similar or in some cases down in 2019 globally. Most importantly, in 2020, the difference is more visible, as NCD dependent death rate went in the USA whereas the global death rate was relatively consistent with the previous year or even went down in some cases. This result suggests that in the developed country the lifestyle has changed, and people are physically less active and largely depend on processed food. In long run, these habits may develop NCD. Also, the environment is another important factor in developing NCDs as it is changing rapidly. On the other hand, most of the global portion consists of 3^rd^ world countries and people support their life with physical work and also have limited opportunities to afford processed food which may contribute to the lower rate of NCD.

Regarding communicable diseases, the population of the USA is less affected than the global population in 2010 and success in reducing it is observed in 2019 globally and in the USA. This success may be the result of the development of the healthcare system and pharmaceutical industry and drug discovery especially antibiotics, particularly for third world countries where the majority of the population lives in an unhygienic environment. However, in the very next year 2020, both the global and the US populations are drastically affected by communicable diseases, particularly COVID-19. In figure 3B, the age group we indicated should have literally less impact by any pathogenic as they are supposed to have a mature and effective immune system to fight against it. A relatively higher effect of COVID-19 in this age group, particularly in the USA where non-communicable health disparities are higher is indicative to compromise immunity due to later one.

In old age, middle age was considered the golden time because of physical fitness, but emerging industrialization has changed that philosophy as a high percentage of the population from this age group is developing NCD at an alarmingly increasing rate. At present most of the global burden of disease is NCDs and imposing a huge health care burden as there is no cure for them. If NCDs continue to increase in this age group, in the new generation will be born with a high risk of developing NCDs and compromise immunity. It is high time to educate young adults about NDC, how it develops and what are the possible consequences. Multiple public health interventions need to device and implement targeting this age group as they are the center for aspects of the community, society, and country. At the same time, they are functioning as parents, children, and decision-makers.

## Conclusion

The major share of the disease burden goes to non-communicable diseases along with the healthcare burden in both global and local scenarios. The analysis finding indicates a higher death rate by non-communicable diseases in the USA while a higher global death rate by a communicable diseases. The study paid more importance to the middle age group for socioeconomic and generational vitality. It highlights the need for continued efforts to address the burden of non-communicable diseases irrespective of the economic status of a country and through developing a multi-level intervention. More importantly, the higher authorities in the world should come together to remodel the global health system to ensure health equity for all levels of people because diseases are also now globalized in this time of globalization and COVID-19 has set an example.

### Implications for public health policy and practice

The study has important implications for public health policy and practice. The findings suggest that efforts to prevent and treat non-communicable diseases should be a top priority for public health policymakers and practitioners. This may require a multi-faceted approach that includes policies and programs aimed at promoting healthy lifestyle habits, improving access to healthcare, and reducing environmental hazards. Additionally, the study highlights the need to address the global variation in disease burden, particularly in low- and middle-income countries where the burden of communicable diseases remains high.

## Data Availability

All data produced in the present study are available upon reasonable request to the authors.

## References

1. MDB [Internet]. [cited 2023 Feb 27]. Available from: https://platform.who.int/mortality

2. Noncommunicable diseases [Internet]. [cited 2023 Feb 27]. Available from: https://www.who.int/health-topics/noncommunicable-diseases#tab=tab_1

3. Noncommunicable diseases [Internet]. [cited 2023 Feb 27]. Available from: https://www.who.int/health-topics/noncommunicable-diseases#tab=tab_1

4. Bennett JE, Stevens GA, Mathers CD, Bonita R, Rehm J, Kruk ME, et al. NCD Countdown 2030: worldwide trends in non-communicable disease mortality and progress towards Sustainable Development Goal target 3.4. The Lancet [Internet]. 2018 Sep 22 [cited 2023 Feb 27];392(10152):1072–88. Available from: http://www.thelancet.com/article/S0140673618319925/fulltext

5. Mortality and global health estimates [Internet]. [cited 2023 Feb 27]. Available from: https://www.who.int/data/gho/data/themes/mortality-and-global-health-estimates

6. CDC - NCHS - National Center for Health Statistics [Internet]. [cited 2023 Feb 27]. Available from: https://www.cdc.gov/nchs/index.htm

7. GLOBAL STATUS REPORT on noncommunicable diseases 2014 “Attaining the nine global noncommunicable diseases targets; a shared responsibility.”

8. Campbell-Lendrum D, Prüss-Ustün A. Climate change, air pollution and noncommunicable diseases. Bull World Health Organ [Internet]. 2019 Feb 2 [cited 2023 Feb 27];97(2):160. Available from: /pmc/articles/PMC6357572/

9. Semenza JC, Rocklöv J, Ebi KL. Climate Change and Cascading Risks from Infectious Disease. Infectious Diseases and Therapy 2022 11:4 [Internet]. 2022 May 19 [cited 2023 Feb 27];11(4):1371–90. Available from: https://link.springer.com/article/10.1007/s40121-022-00647-3

10. di Cesare M, Khang YH, Asaria P, Blakely T, Cowan MJ, Farzadfar F, et al. Inequalities in non-communicable diseases and effective responses. The Lancet [Internet]. 2013 Feb 16 [cited 2023 Feb 27];381(9866):585–97. Available from: http://www.thelancet.com/article/S0140673612618510/fulltext

11. Global Health Estimates [Internet]. [cited 2023 Feb 27]. Available from: https://www.who.int/data/global-health-estimates

12. Gibbons CL, Mangen MJJ, Plass D, Havelaar AH, Brooke RJ, Kramarz P, et al. Measuring underreporting and under-ascertainment in infectious disease datasets: A comparison of methods. BMC Public Health [Internet]. 2014 Feb 11 [cited 2023 Feb 27];14(1):1–17. Available from: https://bmcpublichealth.biomedcentral.com/articles/10.1186/1471-2458-14-147

13. CDC - NCHS - National Center for Health Statistics [Internet]. [cited 2023 Feb 27]. Available from: https://www.cdc.gov/nchs/index.htm

14. Oshakbayev K, Zhankalova Z, Gazaliyeva M, Mustafin K, Bedelbayeva G, Dukenbayeva B, et al. Association between COVID-19 morbidity, mortality, and gross domestic product, overweight/ obesity, non-communicable diseases, vaccination rate: A cross-sectional study. J Infect Public Health [Internet]. 2022 Feb 1 [cited 2023 Feb 27];15(2):255. Available from: /pmc/articles/PMC8759097/

15. Liu Q, Jing W, Liu M, Liu J. Health disparity and mortality trends of infectious diseases in BRICS from 1990 to 2019. J Glob Health [Internet]. 2022 [cited 2023 Feb 27];12:4028. Available from: /pmc/articles/PMC8943566/

